# Large distant deletion disrupts CDKN2A enhancer and predisposes to melanoma

**DOI:** 10.64898/2026.05.07.26352537

**Authors:** Peter A. Johansson, Kelly Brooks, Jane M. Palmer, Vaishnavi Nathan, Mai Xu, Jessica L. Scales, Rebecca Hennessey, Elizabeth A. Holland, Mark Harland, Sharon Hutchison, Pui Ying Chan, Aravind Sankar, Sophie Papiernik, Abigail Dennis, Rohit Thakur, Raj Chari, Helen Schmid, Matthew H. Law, Lisette Curnow, Madeline Howlie, Chloe B. Rodgers, Colette Mustard, Tim D. Bishop, Julia Newton-Bishop, Graham J. Mann, Anne E. Cust, David J. Adams, Kevin M. Brown, Nicholas K. Hayward, Antonia L. Pritchard

**Affiliations:** Cancer Research Program, QIMR Berghofer, Brisbane, QLD, Australia; School of Biomedical Sciences, University of Queensland, Brisbane, QLD, Australia; Division of Cancer Epidemiology and Genetics, National Cancer Institute, National Institutes of Health, Bethesda, MD, USA; Centre for Cancer Research, Westmead Institute for Medical Research, The University of Sydney, Westmead, NSW, Australia; Leeds Institute of Medical Research at St. James’s, University of Leeds, Leeds, UK; Genetics and Immunology Department, University of the Highlands and Islands, Inverness, UK; Experimental Cancer Genetics, Wellcome Trust Sanger Institute, Cambridge, UK; Population Health Program, QIMR Berghofer, Brisbane, QLD, Australia; Faculty of Health, Queensland University of Technology, Brisbane, QLD, Australia; Victorian Clinical Genetics Services, Murdoch Children’s Research Institute, Parkville, VIC, Australia; Melanoma Institute Australia, The University of Sydney, Sydney, NSW, Australia; The Daffodil Centre, The University of Sydney, and Cancer Council NSW, Sydney, NSW, Australia

**Keywords:** Familial melanoma, *CDKN2A*, deletion, non-coding region, upstream, cutaneous melanoma, melanoma

## Abstract

Deleterious *CDKN2A* germline variants account for ∼40% of familial melanoma cases, while rare variants in *CDK4*, *BAP1*, and telomere-maintenance genes collectively attribute ∼10% of familial risk. We sought to identify new high-penetrance susceptibility variants by sequencing 305 melanoma cases from 89 multi-case families negative for known predisposition gene variants. In one family, cutaneous melanoma co-segregated with a rare variant in *DMRTA1* (p.Glu383Gln), located less than 480 kb upstream of *CDKN2A* on chromosome 9. Whole-genome sequencing then revealed an intergenic 234kb deletion that co-segregated with melanoma in 18 out of 21 cases across four generations. Further investigations revealed a further 10 families carrying this deletion, co-segregating with melanoma. The deleted region was predicted to encompass regulatory sequences and to interact with the *CDKN2A* promoter region. Tiled CRISPR inhibition of the predicted enhancer region confirmed interactions between the distant upstream deletion with *CDKN2A* resulting in decreased p16 transcript mRNA expression. Deletion carriers exhibited near-complete loss of p16 mRNA expression from the affected chromosome. This distant non-coding deletion is one of the most common founder variants predisposing to melanoma and reveals a new mechanism controlling p16 expression. Routine screening for this deletion in individuals with perceived high risk of melanoma is warranted.

---

Germline variants in the high penetrance melanoma susceptibility genes *CDKN2A* and *CDK4* account for cutaneous melanoma (CM) development in about 40% of multi-case families, with a further ∼10% accounted for by rare variants in *BAP1* or genes associated with telomere maintenance ^1^. A significant proportion of genetic risk for familial CM therefore remains elusive.

To identify further germline variants associated with risk, we sequenced 305 individuals from 89 multi-case families from the east coast of Australia using a combination of whole exome sequencing (WES) and whole-genome sequencing (WGS), as previously reported ^2–5^. Families were selected for sequencing after screening known high penetrance familial melanoma genes showed no deleterious variants in *CDKN2A* coding, promoter or intronic regions, *CDK4* p.Arg24Cys and p.Arg24His variants, or *BAP1*, *POT1*, *ACD*, *TERF2IP* or *TINF2* loss-of-function variants ^1^. The methodology for sequencing and data processing is given in the online methods. The data for all rare (variant allele frequency (VAF) < 10^−4^) variants resulting in a protein change were selected.

Most notably, a variant in *DMRTA1* (p.Glu383Gln), a gene ∼480 kb upstream of *CDKN2A,* was present in three of four individuals sequenced from a large multi-case family (Extended Data 1) and confirmed by Sanger sequencing. Given the proximity to the well-established familial melanoma gene *CDKN2A*, further members of the family were screened for this variant. A total of eighteen individuals within this family with a history of at least one CM carried the *DMRTA1* p.Glu383Gln variant (Table 1; Extended Data 1). As the initial individuals selected for WES did not carry known pathogenic variants in *CDKN2A*, potential linkage between the *DMRTA1* p.Glu383Gln variant and an unidentified non-coding pathogenic alteration was investigated. Ten family members were analysed by WGS. This revealed a 233.7kb intergenic deletion (chr9:g.22,209,075_22,442,854; hg19), located 234.2 kb upstream of *CDKN2A* (p16 transcript NM_000077.5) and 4 kb downstream of *DMRTA1* (MANE transcript NM_022160.3) (Figure 1), that was present in all individuals carrying the *DMRTA1* p.Glu383Gln variant. The deletion (and the *DMRTA1* p.Glu383Gln variant) was then confirmed to be present in eighteen out of twenty-one individuals in this family affected by CM for whom we had DNA samples or were obligate variant carriers (Figure 1; Table 1). Three individuals with CM who were not deletion carriers were presumed to be phenocopies ^6^ due to high ambient solar UV-radiation in Australia.

**Figure 1:**
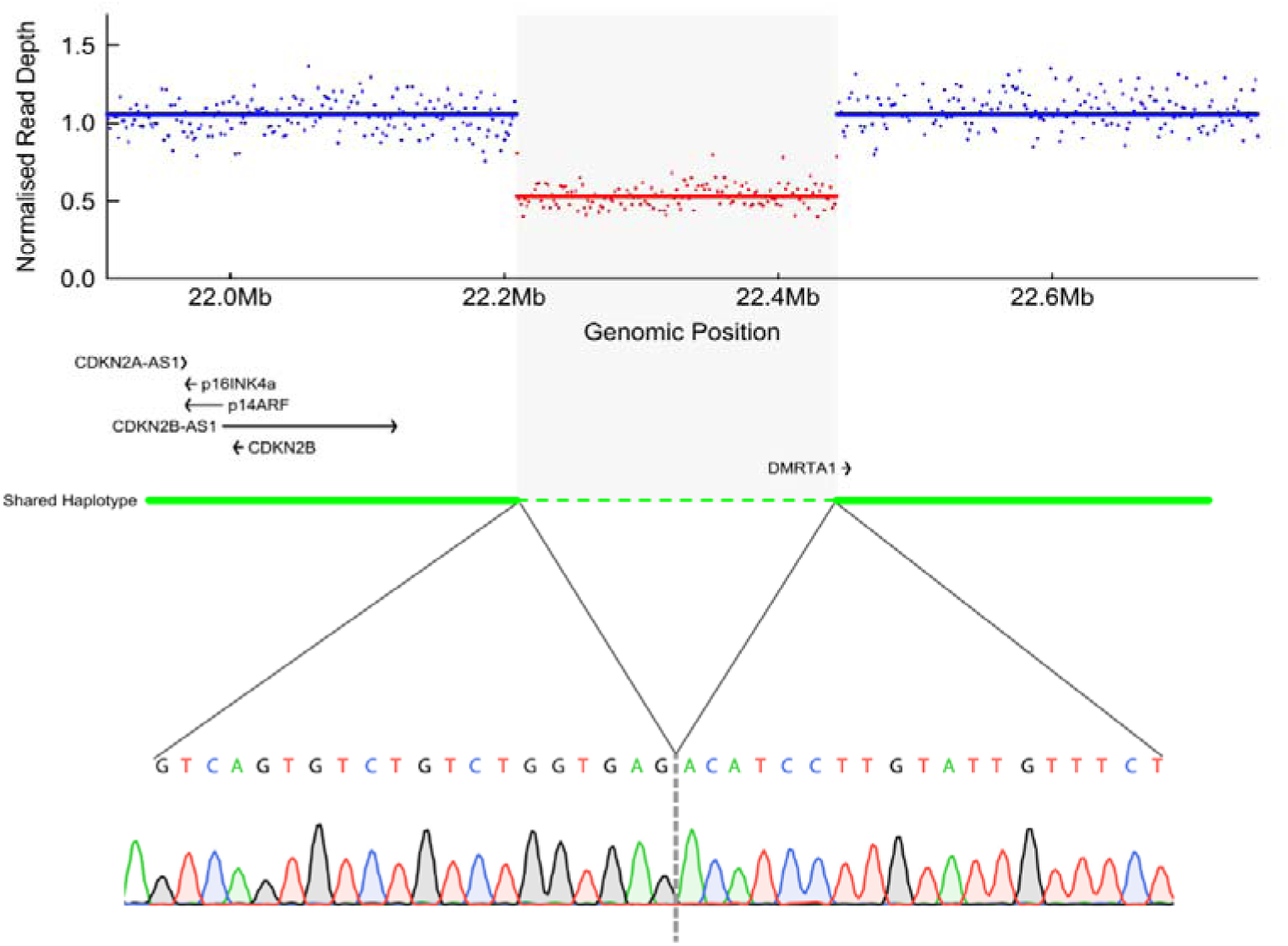
Genomic region on chromosome 9 encompassing the deletion. Whole-genome sequencing read coverage is shown for carrier MELA_1195 from family AUS 1, normalized to the non-carrier MELA_1190 from the same family. Data were binned such that each bin contained 50,000 read bases in the control sample. Each dot represents the ratio of read bases in the carrier relative to the control, normalized by the ratio of total read bases between the two samples. MANE transcripts are shown genes within the region, except for *CDKN2A*, for which both the *p16INK4a* and *p14ARF* transcripts are displayed. The green line indicates the shared haplotype region between families AUS 1 and AUS 3, as inferred from long-read sequencing. The dotted line marks the deletion, which generates a fusion sequence confirmed by Sanger sequencing, shown at the bottom.

**Table 1:** Melanoma prone families carrying the *CDKN2A* proximal deletion.

| Family ID | Number of generations affected with melanoma | Method of identification of deletion | Number of individuals with melanoma | Number of individuals with melanoma who were sequenced or can act as obligate carriers | Number of sequenced melanoma cases or obligate carriers carrying the deletion |
| --- | --- | --- | --- | --- | --- |
| AUS 1 | 4 | WES/WGS | 26 | 21 | 18 |
| AUS 2 | 3 | WGS | 8 | 8 | 8 |
| AUS 3 | 3 | SNP array | 5 | 2 | 2 |
| AUS 4 | 1 | SNP array | 1 | 1 | 1 |
| AUS 5 | 4 | SNP array | 5 | 4 | 4 |
| AUS 6 | 1 | SNP array | 3 | 1 | 1 |
| AUS7 | 3 | WES/WGS | 8 | 6 | 6 |
| AUS 8 | 2 | WES/WGS | 7 | 4 | 4 |
| AUS 9 | 3 | WES/WGS | 4 | 3 | 3 |
| UK 1 | 1 | WES | 1 | 1 | 1 |
| UK 2 | 1 | SNP array | 1 | 1 | 1 |

To determine the prevalence of this *CDKN2A* upstream deletion we investigated three Australian CM cohorts. Firstly, germline WGS data from 303 CM cases in the Australian Melanoma Genome Project (AMGP) ^7,8^ revealed one individual (MELA_0040) carrying the deletion. We were able to obtain DNA samples from four additional family members, who were all diagnosed with CM. Using a PCR assay developed to detect the deletion (online methods and Figure 1) the deletion was also found to be present in all four familial CM cases, as was the *DMRTA1* p.Glu383Gln variant. We then assessed 1611 samples from the Q-MEGA project ^9^ that had been included in genome-wide and polygenic risk association studies for melanoma using single nucleotide polymorphism (SNP) arrays ^10–12^. Copy number variation (CNV) adjacent to *CDKN2A* were assessed using these data. This identified five additional carriers of the deletion, from four families, AUS3, AUS4, AUS5 and AUS6 (Table 1). Further investigation revealed that they all carried the *DMRTA1* p.Glu383Gln variant, and in two families where samples were available from other family members, all but one of the deletion carriers had been diagnosed with CM. Finally, from the Australian Melanoma Family Study (AMFS), a population-based, case-control-family study ^13^, we assessed 596 probands diagnosed with invasive CM before the age of 40. Four probands (0.7%) carried the *DMRTA1* p.Glu383Gln variant and the deletion. One carrier was a member of the index family (AUS 1) who lived in Sydney and took part in both AMFS and the Sydney-based familial melanoma project ^14^ had been found to lack *CDKN2A* exon mutations (online methods). In the other three families (AUS 7, AUS 8, AUS 9) six, four and three CM cases, respectively, had DNA available and were all confirmed to carry the deletion (Table 1). By comparison, ten probands in AMFS were reported to carry other pathogenic p16 variants (1.68%), most commonly p.Glu69Gly (n=3)^15^. The 234kb deletion thus increases the number of probands with p16-affecting variants in this cohort from ten to fourteen, making this deletion a major addition, and the most common p16-affecting variant in this population-based early onset CM cohort.

Given Australia’s predominantly British ancestral background, a British CM cohort ^2^ (cohort 2) was assessed for the presence of the deletion. This identified carriers in two of sixty-six families. One index case (UK1) was identified via harbouring the *DMRTA1* p.Glu383Gln variant and the second index case (UK2) was found to carry the deletion following CNV analysis of SNP array data (Table 1).

To explore whether there was an alternative non-coding variant or structural rearrangement within *CDKN2A* that might have been missed by WES or short-read WGS that was in linkage with the large upstream deletion, two carriers from different families were assessed using long-read sequencing. This confirmed that the 234kb deletion was the only recurrent structural variant in the region.

Long-read sequencing also enabled phasing of SNPs in the region, allowing inference of the haplotype carrying the deletion. This analysis identified a large region (chr9:21,940,588-22,714,093; hg19) shared between AUS families 1 and 3 spanning both *DMRTA1* and *CDKN2A* (Figure 1). Further, the deletion-bearing haplotype included a cytosine (C) at polymorphism rs11515 in the 3’-untranslated region of p16, consistent with the inheritance pattern of genotypes determined by WGS and WES. As guanine (G) is the major allele at rs11515, most deletion-carriers in these families were heterozygous at this site. This confirmation enabled us to use rs11515 as a marker to assess the potential effect of the upstream deletion on *CDKN2A* using allele-specific p16 expression. Analysis of mRNA isolated from circulating white blood cells in deletion carriers (online methods) showed that p16 was expressed almost exclusively from the chromosome without the deletion, indicating that the presence of the deletion resulted in loss of p16 mRNA expression on that chromosome (Figure 2).

**Figure 2:**
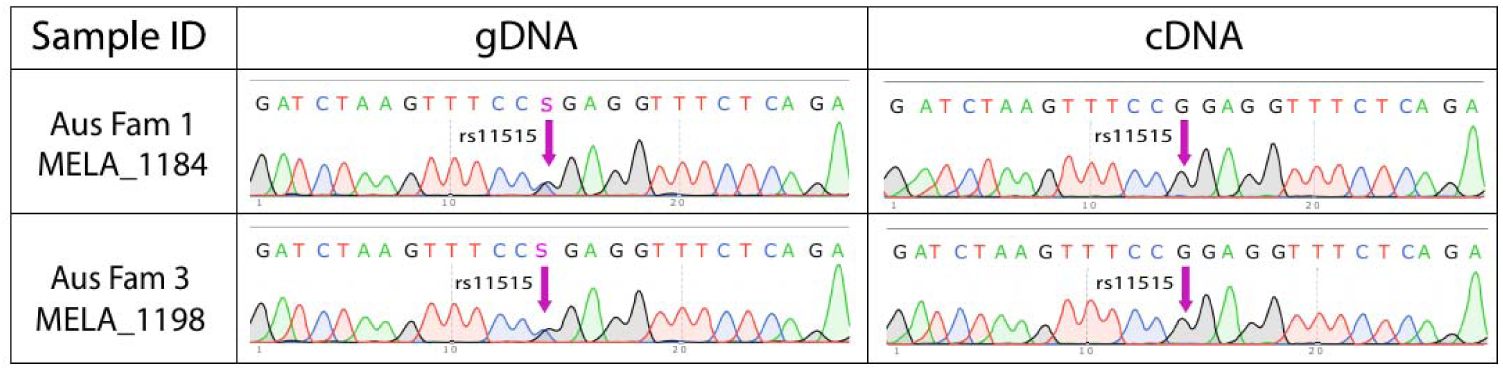
Allele-specific investigation of p16 expression. Presence of both alleles of rs11515, a 3’UTR polymorphism in the *CDKN2A p16* transcript, was confirmed in genomic DNA (gDNA) from two deletion carriers, establishing that rs11515 could be used to tag each chromosome. In mRNA, cDNA analysis showed expression from only the allele originating from the chromosome not bearing the upstream deletion, indicating a loss of p16 expression associated with the deletion.

To explore the potential mechanism by which the deletion alters p16 expression on the same chromosome, data assessing *cis*-chromosomal folding were investigated. The less granular publicly available Hi-C and associated virtual 4C data in the ‘SKCMC’ dataset ^16^ indicated that (i) the deleted region associated with the *CDKN2A* promotor, (ii) *CDKN2A* and approximately half of the deleted region were located within the same topologically associated domain (TAD; ^17^), (iii) the deleted region contained areas predicted to function as enhancers and insulators ^18^ (Extended data 2 A-C). These observations were corroborated by higher-resolution melanocyte capture C experiments ^19^, which showed chromosomal interactions between the deletion and the *CDKN2A* p16 promoter in particular from a region harbouring prominent melanocyte enhancer elements (Extended data 2 D, E). Together, these data suggested a regulatory role for this region of chromosome 9 that is removed in the deletion, resulting in the loss of p16 expression observed in deletion carriers.

The two putative melanocyte enhancer regions identified ^18–20^ were then targeted by CRISPR-inhibition (CRISPRi) using two pools of small guide RNAs (sgRNAs) in immortalized human melanocytes (Figure 3, online methods). Relative to infections with two non-targeting guides, targeting of Enhancer 1 resulted in significant reduction of both the p14 and p16 transcripts of *CDKN2A* (p14 *P_NTC1_* = 1.08 × 10^−4^ and *P_NTC2_* = 1.75 × 10^−5^, 2.37- and 2.37-fold decrease, relative to NTC1 and NTC2 respectively; p16 *P_NTC_1* = 3.28 × 10^−5^ and P_NTC2_ = 1.24 × 10^−3^, 1.87- and 2.10-fold decrease), while targeting of Enhancer 2 did not result in significant alterations (Figure 3 B, C). Consistent with a potential larger role of these regions in *ci*s-regulation of 9p21 genes, significant inhibition of *CDKN2B* when targeting Enhancer 1 (*P_NTC1_* = 6.24 × 10^−4^ and *P_NTC2_* = 1.00 × 10^−4^, 2.39- and 3.02-fold decrease; Figure 3D) and subtle inhibition for Enhancer 2 (*P_NTC1_* = 0.12 and *P_NTC2_* = 8.58 × 10^−3^, 1.29- and 1.63-fold decrease) were observed. Individual sgRNAs targeting a prominent region of open chromatin within Enhancer 1 (Figure 3E) showed significant reduction of p16 in immortalized melanocytes, the most potent of which reduced p16 levels 2.56- and 2.66-fold relative to the two respective non-targeting guides (*P_NTC1_* = 3.33 × 10^−7^ and *P_NTC2_* 4.60 × 10^−11^; Figure 3F). Likewise, the same sgRNA resulted in 3.35- and 3.16-fold knockdown of p14 relative to non-targeting guides (*P_NTC1_* = 3.89 × 10^−6^ and *P_NTC2_* 2.11 ×10^−6^; Figure 3G;). Consistent with the pooled data, significant repression of *CDKN2B* was also observed (Figure 3G).

**Figure 3.**
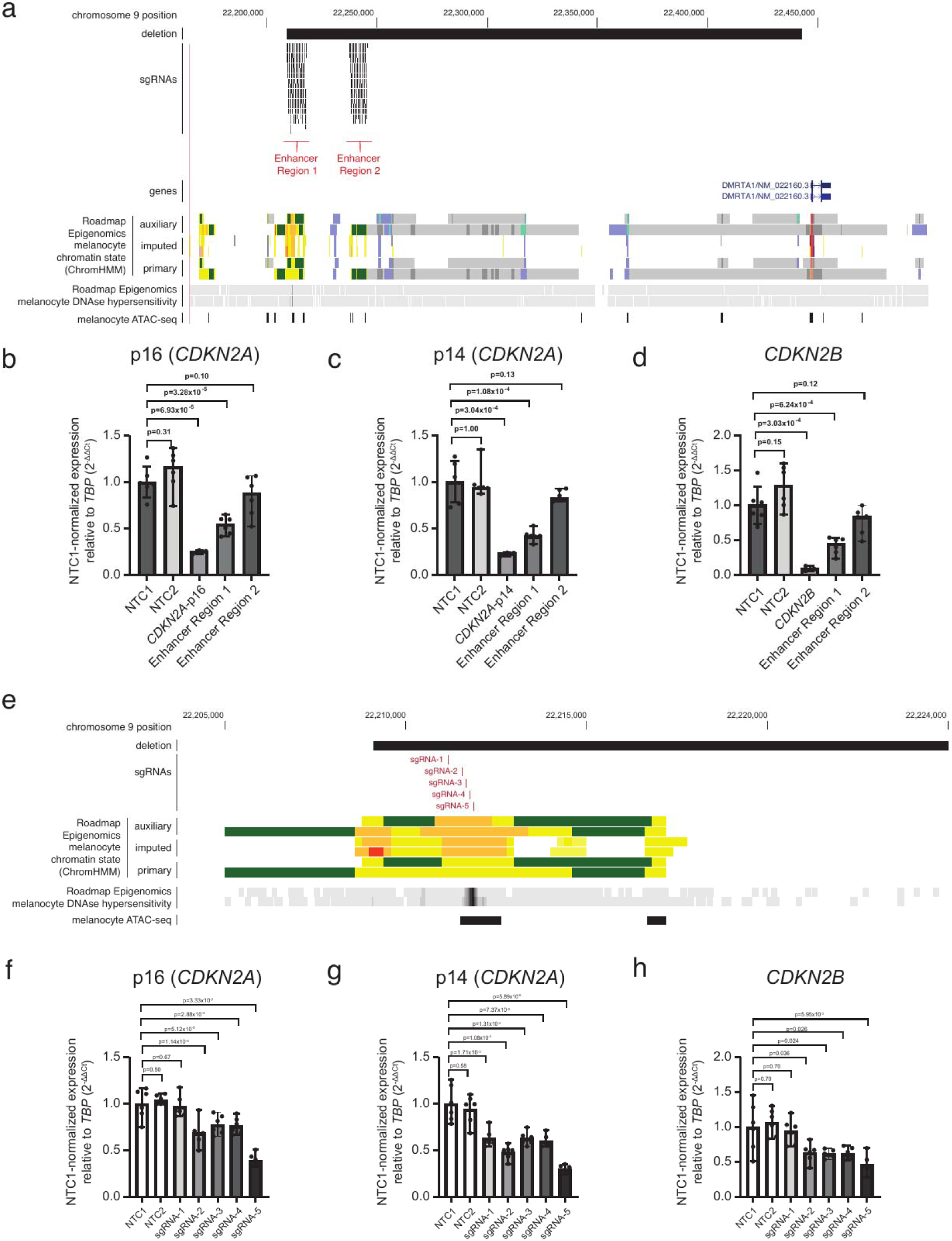
CRISPR-inhibition of a putative melanocyte enhancer region located within the 9p21 deletion inhibited expression of the p16 and p14 transcripts of *CDKN2A*. Pools of small guide RNAs (sgRNAs) were designed to respectively target each of two predicted enhancer regions located within the telomeric end of the deletion. (**A**) Melanocyte chromatin state (ChromHMM) data from Roadmap Epigenomics was visualised from the UCSC Genome Browser; orange, yellow, and yellow-green represents enhancers, reds represent promoters. Melanocyte DNA accessibility from DNAse I hypersensitivity sequencing data from RoadMap Epigenomics, as well as previously published ATAC-seq data were also visualised. (**B-D**) sgRNA pools were expressed in immortalized melanocytes via a lentivirus for dual expression of sgRNAs and dCas9-KRAB, with expression of 9p21 genes assessed by TaqMan qRT-PCR assays. Expression levels were calculated using the 2-ΔCt method with *TBP* as the housekeeping control. Enhancer sgRNA pools were compared to two non-targeting control sgRNAs (NTC1 and NTC2), as well as two pooled sgRNA guides targeting the promoters of p14, p16, and *CDKN2B* as positive controls. Displayed *P*-values were calculated based on 2-ΔCt values from biological replicates relative to NTC1, each with three technical replicates using unpaired two-tailed t-test for comparison to each of the two non-targeting guides for guides targeting enhancers and Welch’s-corrected unpaired, two-tailed t-test for positive controls. (**E**) Five individual sgRNAs were designed to target a region of open chromatin in melanocytes contained within Enhancer 1. (**F-G**) Individual sgRNAs were expressed in immortalized melanocytes and expression of 9p21 genes assessed by TaqMan qRT-PCR assays as described above. For both pooled and individual CRISPRi experiments, statistics are presented for comparison to NTC1 only.

Finally, tumour WGS data were available from two individuals carrying the upstream deletion ^7,21^. The data from the AMGP sample showed a somatic p.Pro114Leu mutation, deleterious to p16 function ^22^ and the available tumour data from a UK sample showed a somatic deletion over *CDKN2A* that affected exons 2 and 3 of p14 and p16. The WGS data did not, however, allow phasing of these tumour-specific mutations to determine whether they occurred on the same chromosome as the germline deletion or resulted in biallelic loss of p16, though the latter seems likely.

Here we describe a 234kb deletion upstream of the key familial melanoma gene *CDKN2A* that co-segregates with cutaneous melanoma in nine Australian and two UK melanoma-prone families and profoundly reduces p16 expression. In the general population, *DMRTA1* p.Glu383Gln was present in 90 individuals in the gnomAD collection (n=807,019 samples) ^23^, 89 of whom had European ancestry and one with ambiguous ancestry, suggesting this variant arose in the European population (non-Finnish European VAF, 7×10^−5^). Three individuals in the gnomAD structural variant collection ^24^ (n= 63,046 samples) carried the 234kb deletion, all of whom were non-Finnish-European (VAF 5×10^−5^). In the Database of Genome Variants (DGV), for which a single study using genome-wide SNP analysis of 48,669 individuals ^25^ was the main contributor, an overlapping deletion was present once (nsv1016067; chr9:22,210,874-22,432,654, hg19). There were additionally deletion structural variants that overlapped with the deleted region to a lesser degree (nsv1015364; nsv1023972). No cancer phenotype data are available in gnomAD and DGV. The deletion we identified here, associated with loss of p16 expression and increased risk of familial CM, is uncommon in the general population and represents a common founder pathogenic variant in melanoma families of European ancestry. Together, the chromosome looping and CRISPRi data provided evidence of a prominent enhancer region contained within the deleted region influencing *CDKN2A*, giving mechanistic insight into the p16 expression loss in deletion carriers and the increased melanoma predisposition observed in the families. This finding therefore substantially extends our understanding of melanoma risk associated with chromosome band 9p21, by providing a new method by which p16 is controlled.

In conjunction with a companion report in this edition of the journal, describing a different founder deletion spanning an overlapping region of chromosome 9 in Italian CM families, our findings establish noncoding structural variation at 9p21 as a significant and recurrent melanoma predisposition mechanism with broad implications. These findings support a substantial increased proportion of familial melanoma being explained by alteration to p16 function through a novel mechanism of expression control. More generally, they highlight the potential to identify additional cancer predisposition variants in noncoding regions some distance from the genes they regulate and suggest inclusion of this deletion in routine *CDKN2A* genetic testing for melanoma risk assessment.

---

**Extended Data 1: Pedigree of family AUS 1 carrying a large deletion upstream of CDKN2A that segregates with melanoma.**

Available upon reasonable request to authors

**Extended Data 2:**
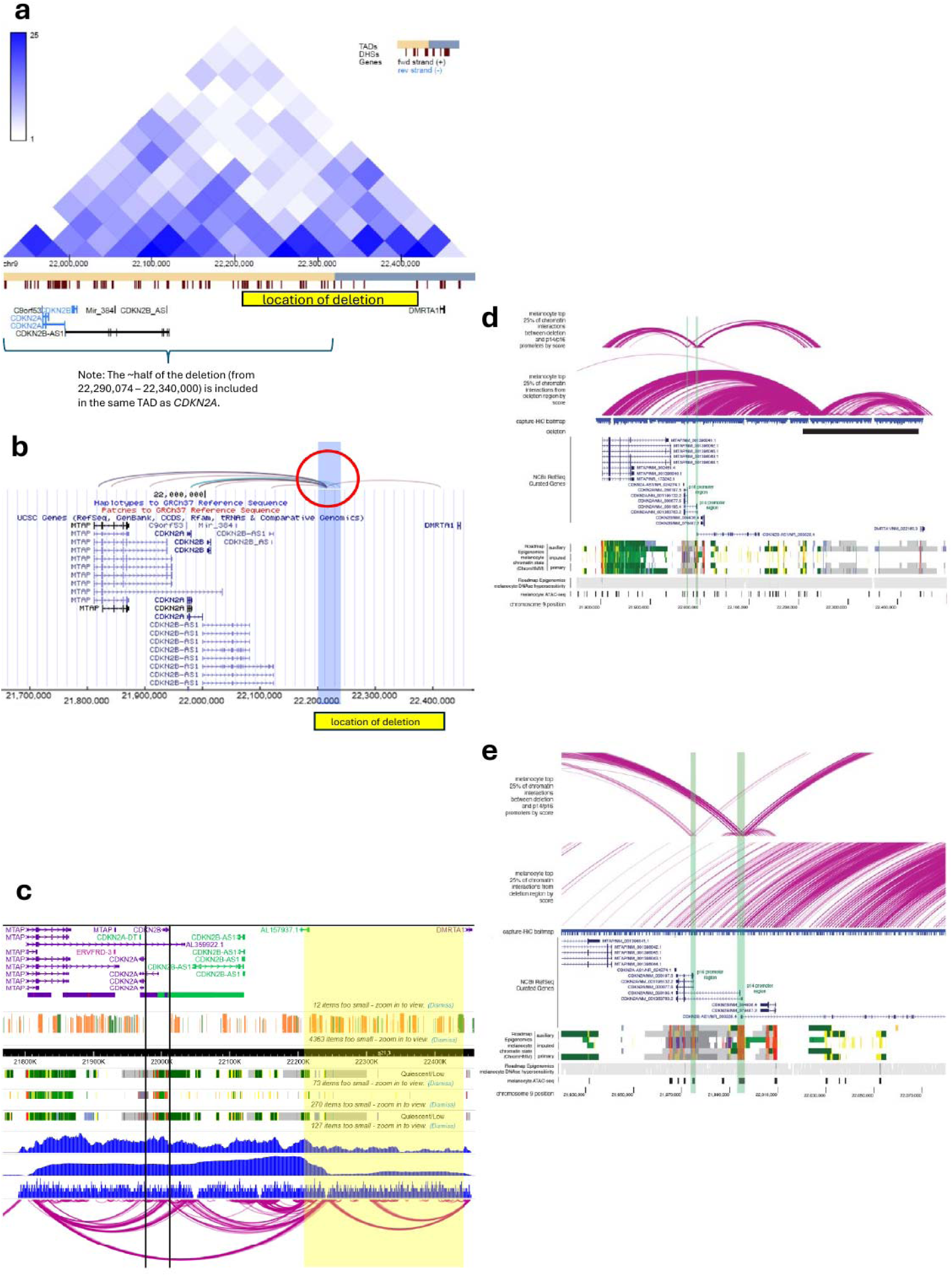
Chromatin interactions with the 9p21 deletion region in melanocytes. Chromatin interactions from human primary melanocytes via a custom capture-HiC is plotted for (A) the wider 9p21 region (chr9:2175000-22500000, hg19) and (B) over CDKN2A (chr9:21720000-22080000). Chromatin loops have been filtered to show only the top 25% of interactions by score. Capture-baits are annotated in blue, with the deletion region represented in black. Plotted genes are NCBI RefSeq Curated genes, with exception of CDKN2B-AS1, for which only a representative (MANE) transcript is shown. Melanocyte chromatin state (ChromHMM) data from Roadmap Epigenomics was visualised from the UCSC Genome Browser; orange, yellow, and yellow-green represents enhancers, reds represent promoters. Melanocyte DNA accessibility is shown from DNAse I hypersensitivity sequencing data from RoadMap Epigenomics, as well as previously published ATAC-seq data. Annotated promoter regions for p16 and p14 are highlighted in green in both panels.

## Methods

### Familial melanoma sequencing

#### Queensland Familial Melanoma Study

All participants gave written informed consent for participation (ethical approval ID P452) and were wild-type for germline *CDKN2A* and *CDK4* mutations. Study participants were selected from those individuals ascertained as part of the Q-MEGA project ^1^, or study participants were recruited following referral from clinicians. Q-MEGA was a population-based study investigating the link between genetics and environment in melanoma development and consists of four study samples: The Queensland Study of Childhood Melanoma (*n*=101); The study of Melanoma in Adolescents (*n*=298); The Study of Men over 50 (*n*=178); and the Queensland Familial Melanoma Project (QFMP; *n*=1897) ^2^. All participants provided detailed information on personal and family cancer history; additional family members (both cancer affected and unaffected) were invited to participate. Whole genome (WGS) or exome (WES) sequencing was performed on high-density melanoma affected families, resulting in between 2 and 5 melanoma cases being sequenced using whole exome sequencing (WES) or whole genome sequencing (WGS) per family (totalling 305 participants with a history of at least one melanoma, from 89 families).

Sequencing was performed by Macrogen (South Korea) on the Illumina HiSeq 2000 platform, with mean coverage of 60 to 96x (WES) and 30-38x (WGS). Using the Burrows-Wheeler Aligner, the sequence output was aligned to the 1000 Genome Project human reference genome (humanG1Kv37) ^3^. Duplicate reads were marked with Picard MarkDuplicates and alignments refined using GATK ApplyBQSR and BaseRecalibrator ^4^. Small variants were called with GATK HaplotypeCaller and filtered to remove low-quality variants defined to have phred-scaled likelihood (PL) for reference genotype < 70, fewer than three variant-supporting reads, significant read position bias (Mann-Whitney *P* < 0.0001), or mapping quality bias (Mann-Whitney *P* < 0.0001). Variants were annotated using Ensembl Variant Effect Predictor ^5^ and further annotated with gnomAD v4.1 allele frequencies ^6^ using an in-house-tool. Large structural variants were called in WGS data using janda (https://janda.sourceforge.net/).

#### Australian Melanoma Family Study

597 probands, diagnosed with their first CM between 1 July 2000 and 31 December 2002 and before the age of 40 years of age, were ascertained from the Australian Melanoma Family Study ^7^. Some probands, or their relatives, had previously been recruited to a Sydney-based familial melanoma project initiated by the Sydney Melanoma Unit ^8^.

#### Q-Mega CNV analysis

Data from the SNP arrays performed on samples collected from participants in the Q-MEGA project ^1^ for genome wide association studies ^9,10^ and polygenic risk score analyses ^11^ were analysed using GenomeStudio 2.0 (Illumina) using the CNV analysis plugin (cnvPartition v.3.2.0). Settings were provided to ensure that only robust regions of loss and gain were called (minimum probe count n=10; minimum homozygous region = 1Mb; Confidence threshold = 35).

**Online Methods Table 1:**
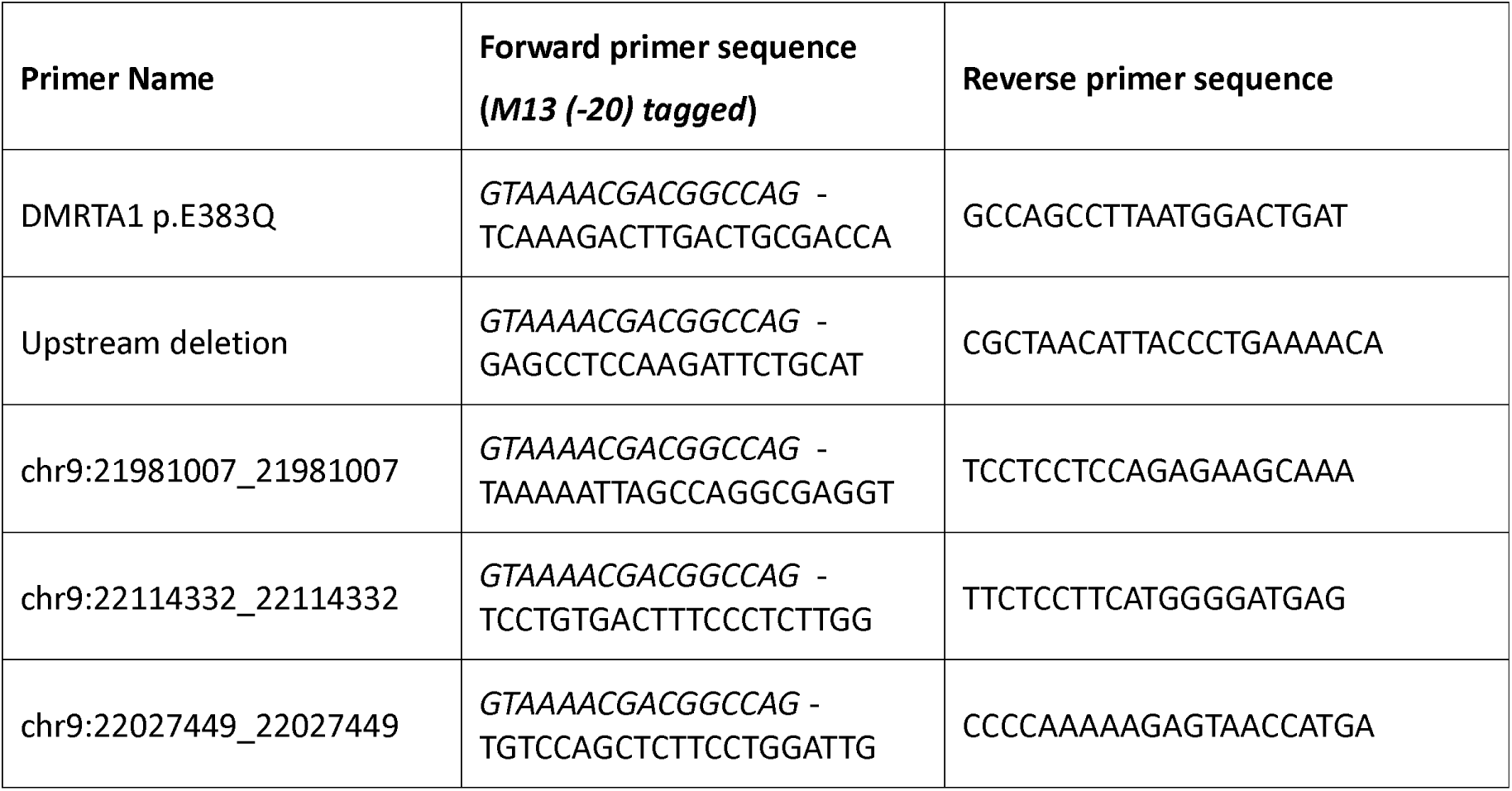
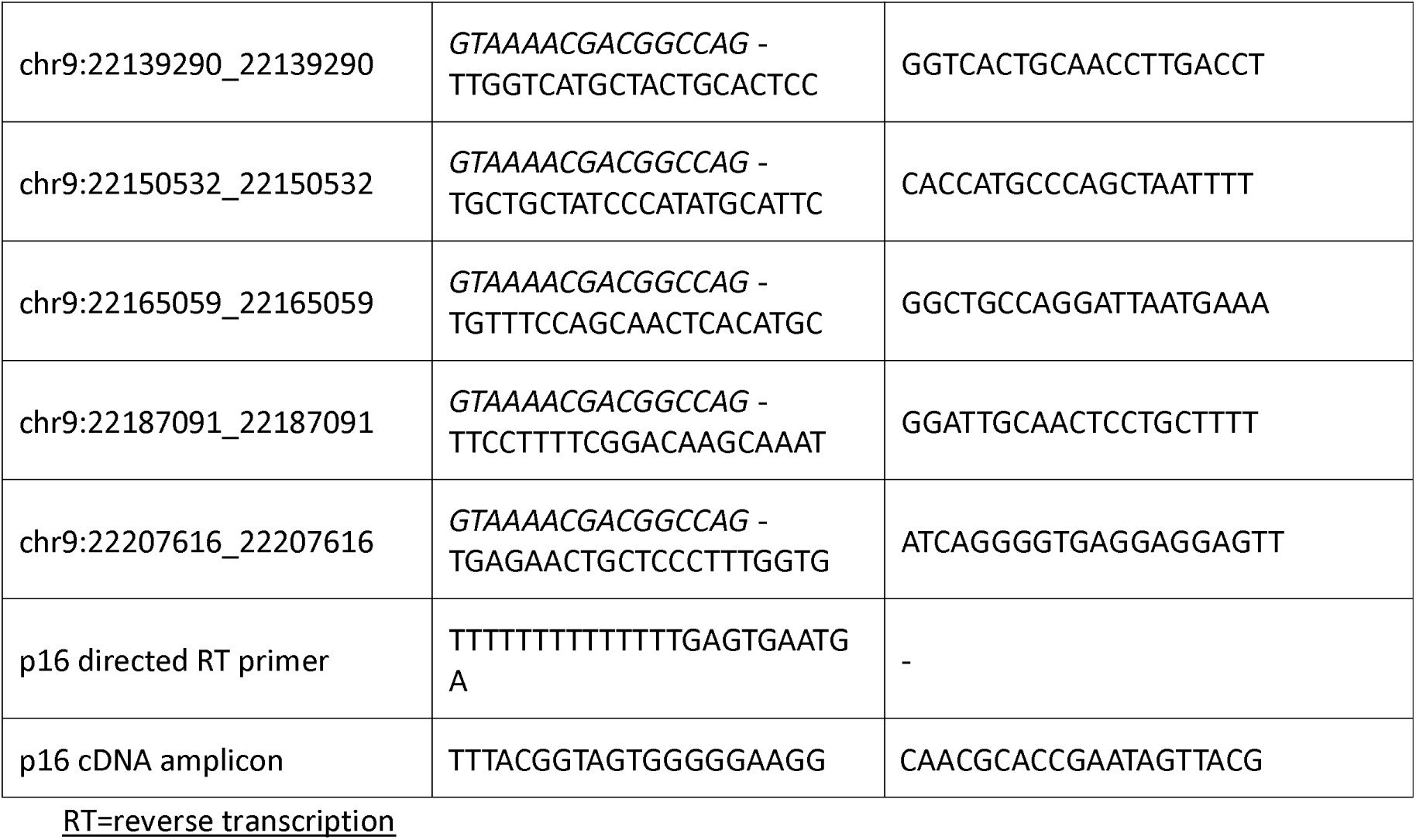

#### Sanger sequencing

PCR was performed using GoTaq® Master Mix (Promega), following the manufacturer’s protocol; primers are detailed in Online Methods Table 1. PCR products were visualised using agarose gel electrophoresis, after which Sanger sequencing was carried out using the BigDye™ Direct Cycle Sequencing Kit (Applied Biosystems), the AMPure XP beads (Agencourt) for PCR cleanup and the CleanSEQ Dye Terminator Removal beads (Agencourt) for sequencing reaction cleanup. After sequencing, the resulting chromatograms were visualised using SnapGene.

#### PacBio sequencing

Long read PacBio sequencing was performed at the central sequencing services at the NIH Molecular Genomics Sequencing Core Facility. Consensus reads were generated with pb_css and aligned to the reference genome with pbmm2. Small variants were identified using DeepVariant v0.9.0 and GATK HaplotypeCaller v4.2.2.0. Structural variants were identified by cuteSV v1.0.13 and pbsv v2.6.2. SNVs were phased with the large deletion using an in-house tool.

#### Analysis of p14 and p16 mRNA expression in the blood of deletion and non-deletion carriers

Blood was collected by venepuncture into Paxgene Blood RNA Tubes (Qiagen) and processed within 2 weeks of collection for mRNA isolation using the PAXgene Blood RNA Kit (Qiagen). The mRNA was reverse-transcribed to cDNA using a semi-specific *CDKN2A* primer (Online methods Table 1). Amplicons were created using primers specific for the p16 sequence (Online methods Table 1) and Sanger sequencing was performed using reverse primers and standard protocols (Applied Biosystems) based on the original principle ^12^ to ensure the 3’UTR SNV (rs11515 VAF = 0.86) was well into the chromatogram trace. Genomic DNA (gDNA) was sequenced using an amplicon that spanned the same 3’UTR SNV, using the reverse primers previously detailed, but with a forward primer that sat in intron 2 (Online methods Table 1). Sanger sequencing results were visualised using SnapGene. The exon 2/exon 3 border was assessed to ensure that no gDNA had been amplified.

#### Promoter capture Hi-C on Melanocytes

Promoter capture Hi-C is a method that enriches a standard Hi-C library to specifically identify which regions of the genome physically interact with promoters to enable genome-wide detection of distal promoter-interacting regions ^13^. The protocol used to derive the promoter capture Hi-C data in melanocytes used in this study was recently described in detail by Thakur et al ^14^. Data from these experiments were used for specific analysis of the CDKN2A -> DMRTA1 region for chromosomal regional interactions.

Briefly, capture HiC on primary neonatal human melanocyte cultures isolated from discarded foreskin tissue of healthy newborn males from five unrelated donors were assessed as a part of a larger study of 54 common melanoma risk loci ^14^. In the chromosome 9 region around *CDKN2A*, capture baits were tiled across restriction fragments covering a region from *MTAP* to *DMRTA1* (chr9:21,790,755–22,452,478, hg19). Given that all interactions in this region were bait-to-bait interactions, visualisations were filtered to retain only the highest scored bait-to-bait interactions in the region (CHiCAGO score).

#### Cell culture

C283T immortalized melanocytes ^15^ were cultured in Gibco Cascade Biologics M254 (Invitrogen, Cat# M-254-500) supplemented with HMGS-2 (Invitrogen, Cat#S0165), grown at 37°C with 5% CO_2_. for Capture Hi-C experiments. For CRISPRi experiments, cells were grown in Dermal Cell Basal Medium (American Type Culture Collection (ATCC), PCS-200-030) supplemented with Melanocyte Growth Kit (ATCC, PCS-200-041) and 1% amphotericin-B/penicillin/streptomycin (Quality Biological, 120-096-711). All cells tested negative for mycoplasma via the MycoAlert PLUS Mycoplasma Detection Kit (Lonza, Cat# LT07-710).

#### CRISPR inhibition

Enhancer regions were defined via Hi-C, 4C, Capture C and visual inspection of ChromHMM data from RoadMap Epigenomics data derived from two neonatal human melanocyte cultures ^14,16,17^. They were defined as: Enhancer 1: chr9:22209074-22217765; Enhancer 2: chr9: 22,237,600-22,245,400 (coordinates are hg19). Two sgRNA pools (Enhancer 1 (E1) and Enhancer 2 (E2)) were designed using sgRNA Scorer 2.0 ^18^. On average, density for these regions was two sgRNAs per 100 nucleotides; E1 was covered by 176 guides; E2 was covered by 157 guides. Oligo pools were synthesized by Twist Biosciences with appropriate flanking sequences for isothermal assembly. PCR product was generated from the oligo pool and subsequently cloned into plasmid pRC1088 which was digested with BsmBI using the NEB HiFi Builder. The assembled product was column purified and subsequently electroporated into Lucigen Endura electrocompetent cells. Cells were then recovered with Super Optimal Broth with catabolite repression (SOC) and plated across 10 - 15 cm LB-Agar plates with Carbenicillin. Plates were grown for ∼20 hrs at 30°C and then bacterial colonies were scraped and pelleted and plasmid DNA was isolated using the ThermoFisher Genejet Endo-free Maxiprep kit. pRC1088 is a modified version of Addgene-71237 ^19^ where green fluorescent protein (GFP) was replaced with puromycin. Addgene-7137 (pLV hU6-sgRNA hUbC-dCas9-KRAB-T2a-GFP) was a gift from Charles Gersbach (Addgene plasmid # 71237 ; http://n2t.net/addgene:71237 ; RRID:Addgene_71237).

Illumina sequencing was performed once the cloned plasmid library was generated to look at the sgRNA counts and then plot the cumulative proportion to look for any skew. Two non-targeting gRNAs (NTC1, NTC2) were separately used as negative controls; pools of two gRNAs targeting each of the *CDKN2A*-p14, *CDKN2A*-p16, and *CDKN2B* promoter regions were used as positive controls. sgRNA sequences and locations are listed in Online Methods Table 2 and enhancer-targeting sgRNAs are visualized in Figure 3. All sgRNAs were cloned into pRC1088 (Addgene #71237), a dual vector co-expressing dCas9-KRAB and gRNA. Virus for expression of various sequences were first titrated using an HIV p24 ELISA kit from ZeptoMerix (RETROTEK™ HIV Type 1 p24 Antigen ELISA, Cat #080111), which showed a similar titer for different gRNA constructs. Based on the measured titre, we further tested cell viability after infection and puromycin selection using a virus amount of ∼100TU per cell (or MOI=100). Virus targeting ∼80% survival of cells was used. Immortalized human melanocyte C283T cells were infected with lentiviral particles encoding individual sgRNAs (non-targeting guides) or respective sgRNA pools (Enhancer 1, Enhancer 2, positive control genes). At 24 hours after infection, 1.2 μg/mL of puromycin was added for selection. After two days of puromycin selection, puromycin was removed, and cells were harvested for RNA isolation. Six infections were performed for enhancer-targeting sgRNA pools as well as non-targeting sgRNAs; three were performed for positive control genes.

mRNA levels of *p14*, *p16* and *CDKN2B* were measured by TaqMan qPCR assay and normalized to *TBP* levels (Thermo Scientific; CDKN2A-p16: Hs02902543-mH; CDKN2A-p14: Hs00924091-m1; CDKN2B: Hs00793225-m1; TBP: Hs00427620-m1). Assays were run with standard time conditions (1.6°C/s for: 2 min at 50°C, 10 min at 95°C, and 40 cycles of [15 s at 95°C, 1 min at 60°C]) on an Applied Biosystems QuantStudio 7Flex Real-Time PCR System with QuantStudio Real-Time PCR Software (v1.7.2). A 10 μl or 20 μl reaction was set up using 50 ng cDNA with 1X TaqMan Fast Advanced Master Mix (Thermo Fisher Scientific, Cat# 4444964) and 1X TaqMan gene expression assay probes. All samples were run in a 384-well plate. Gene expression was determined using the ΔCt method. Expression levels were calculated using the 2^−ΔCt^ method with TBP (VIC-labeled) as the housekeeping control. *P*-values were calculated based on 2^−ΔCt^ values from biological replicates, each with three technical replicates using a two-tailed t-test for comparison to each of the two non-targeting guides for guides targeting enhancers and Welch’s-corrected unpaired, two-tailed t-test for positive controls.

For individual gRNA validation, gRNA_1 to gRNA_5 were cloned into pXPR-050 vector (gift from John Doench and David Root, Addgene plasmid #96925; RRID: Addgene_96925) and dCas9-KRAB expressing C283T cells were infected with viral particles expressing each individual gRNA. Cells were then treated with puromycin and processed for RNA purification and TaqMan assays, same as described in the pooled gRNAs experiment.

**Online Methods Table 2:**
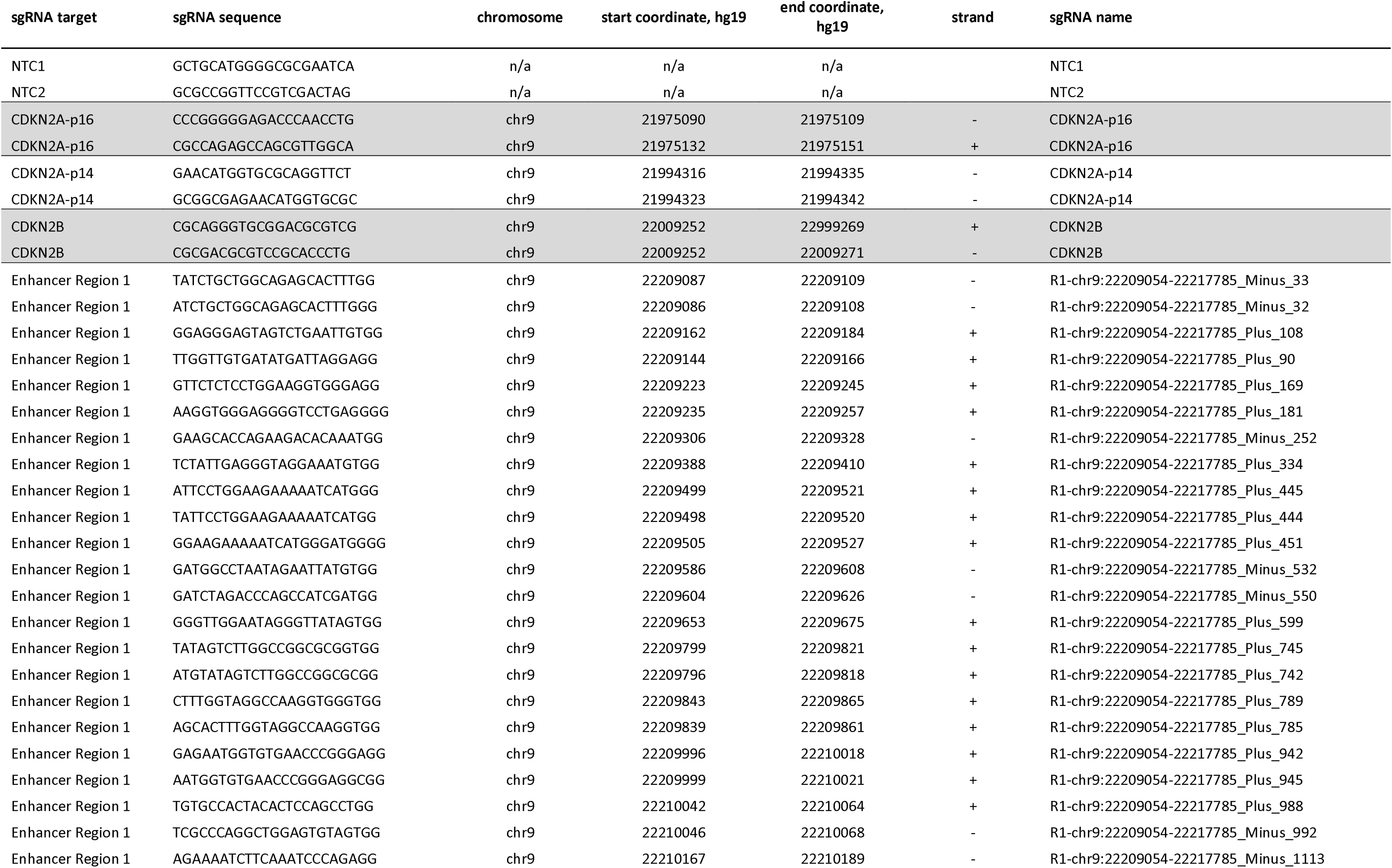
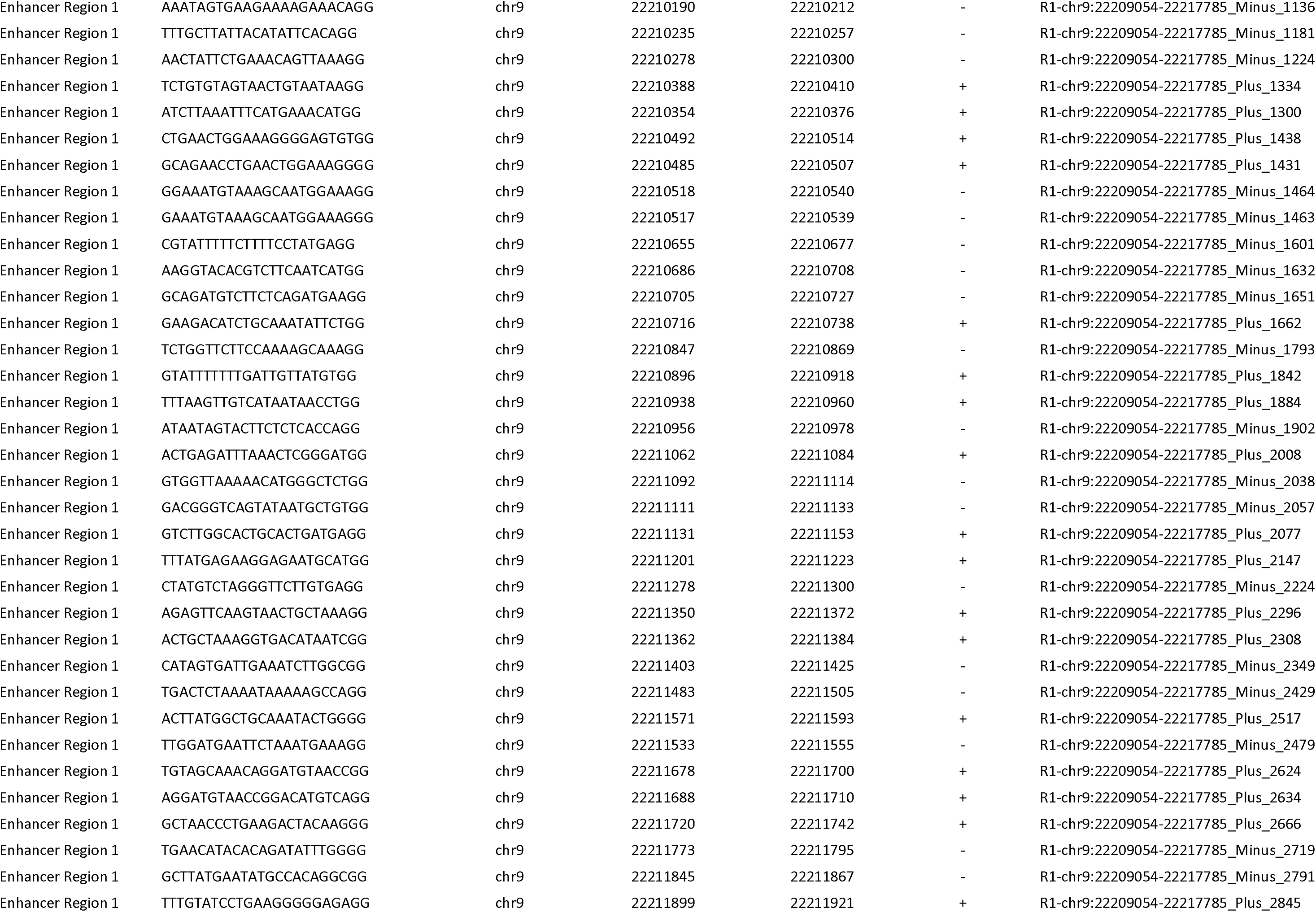
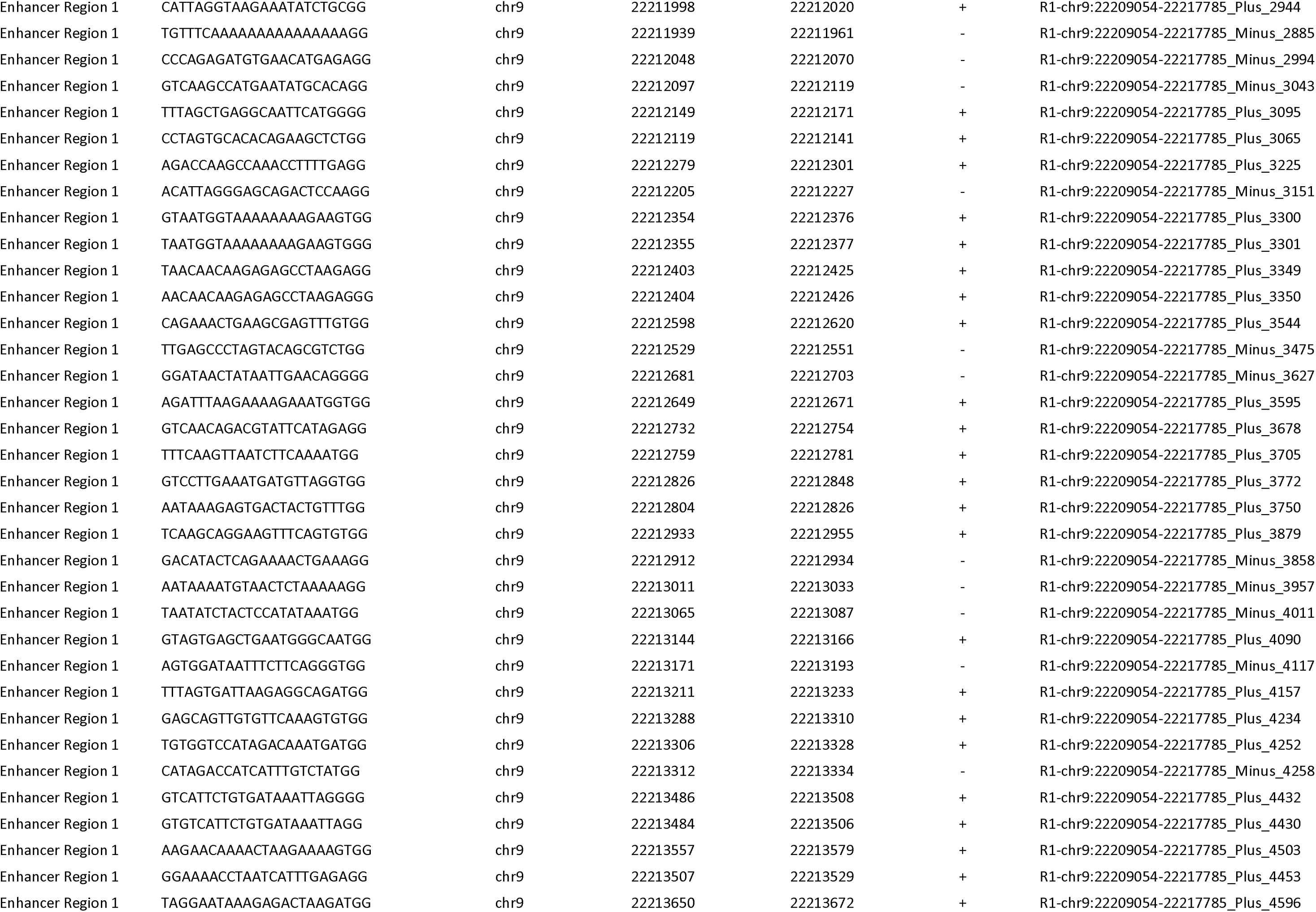
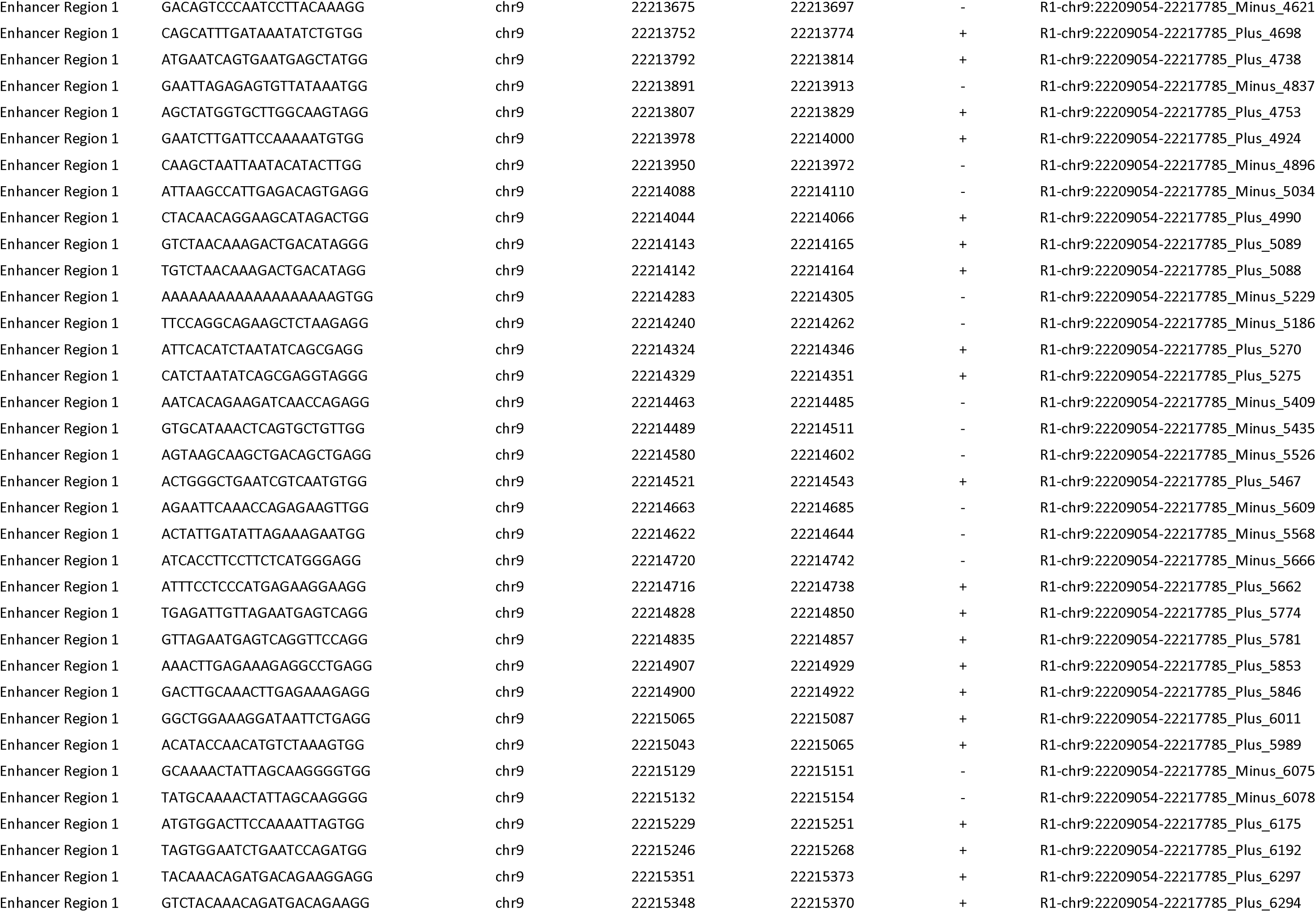
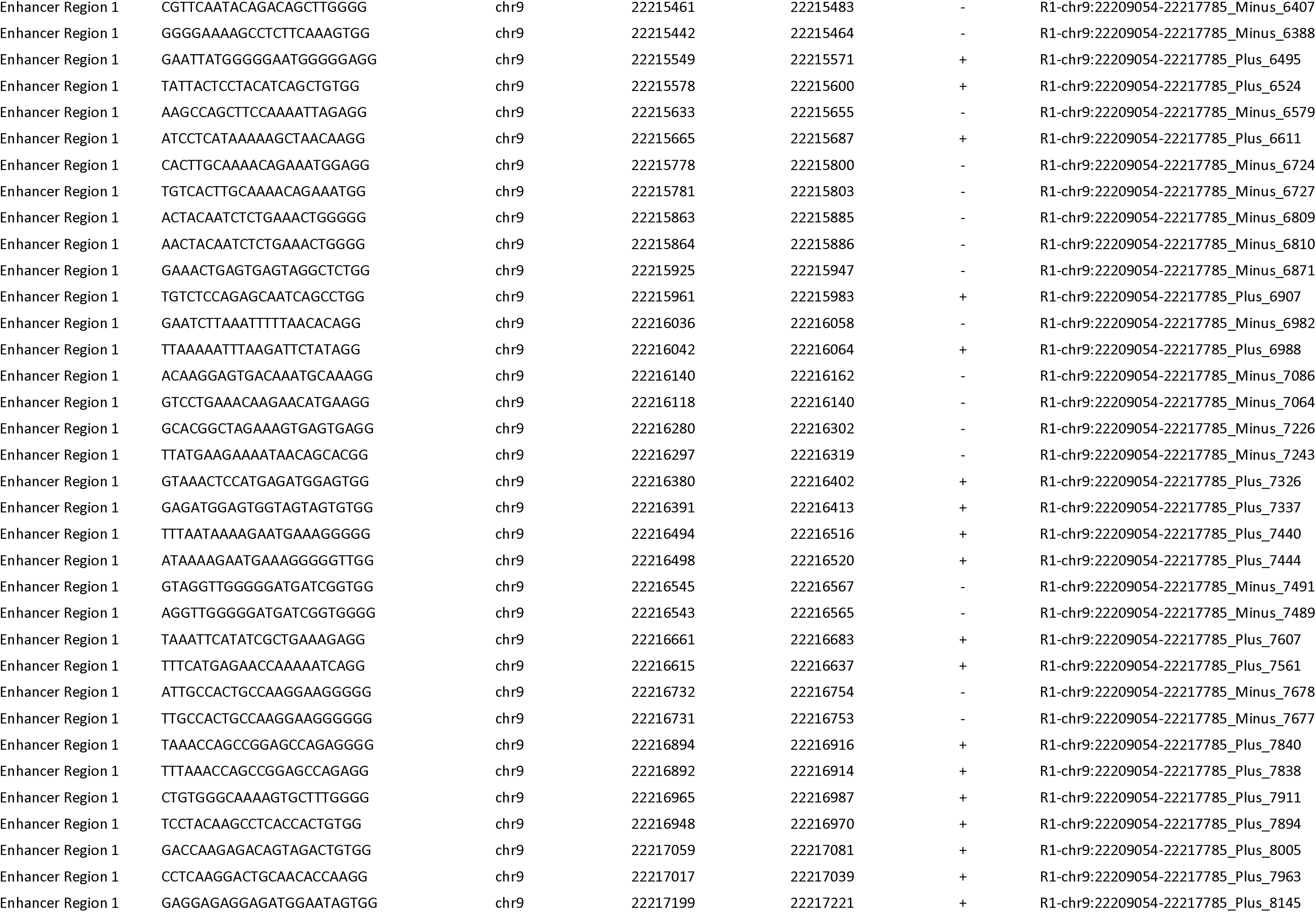
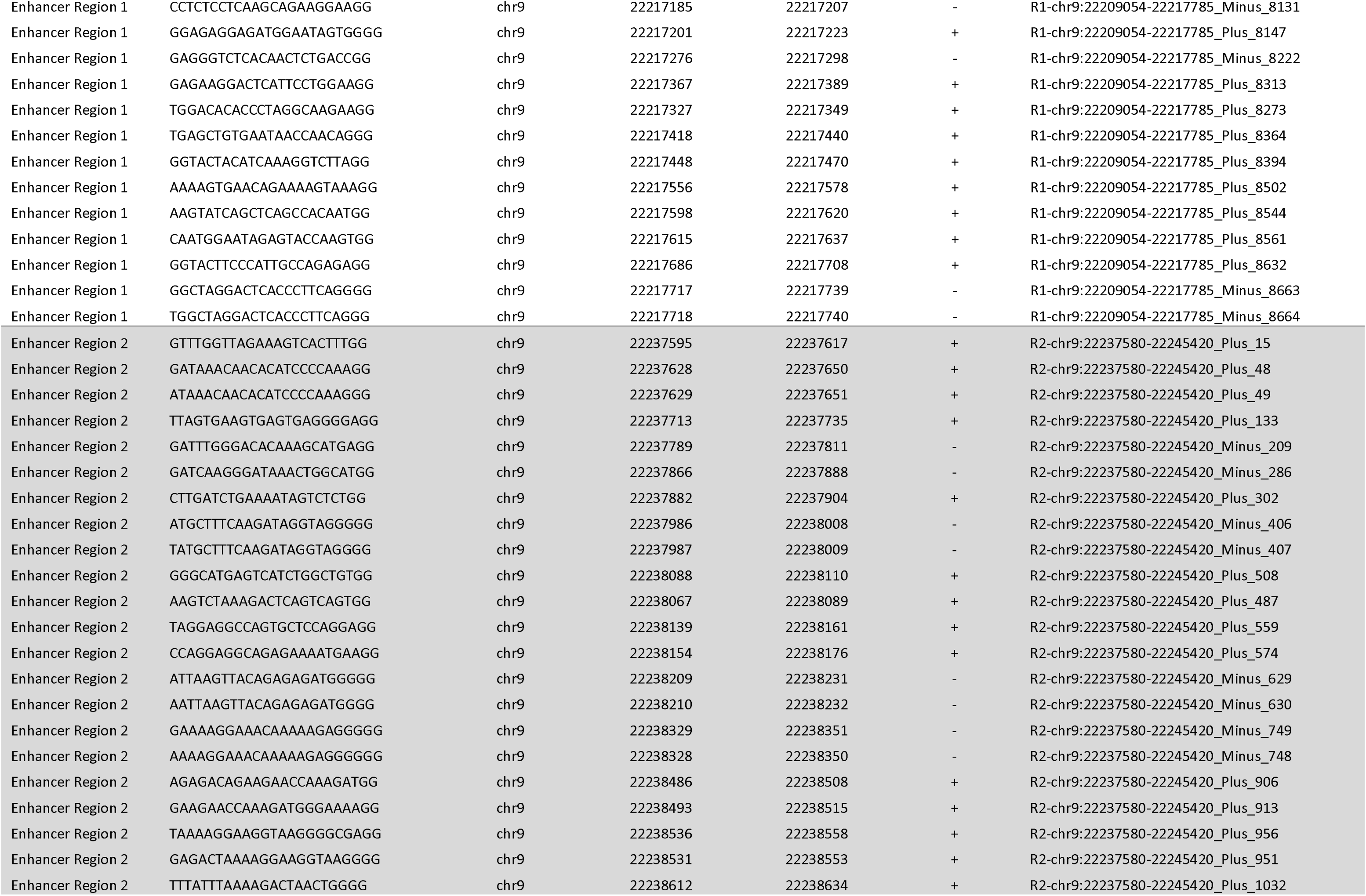
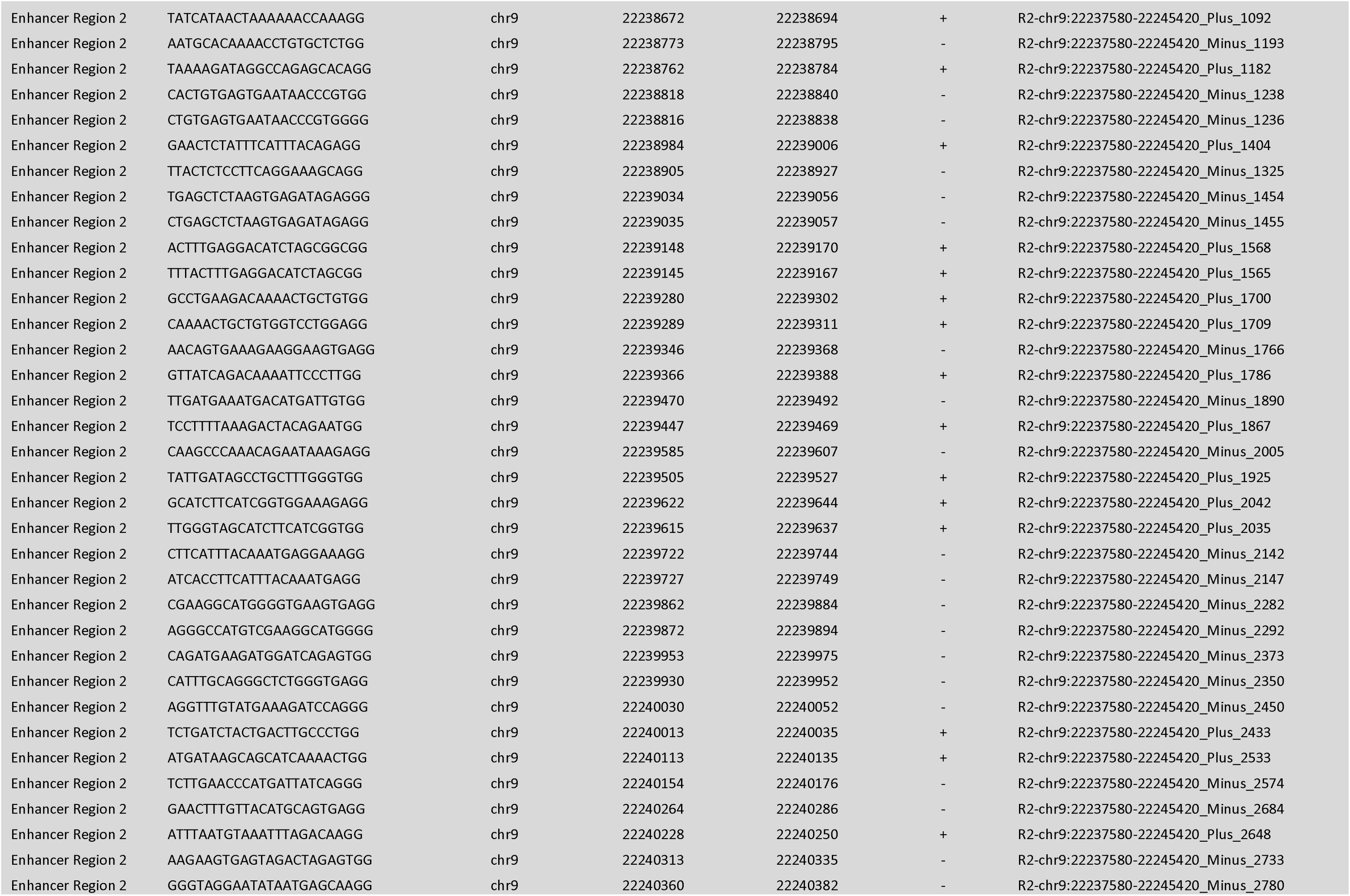
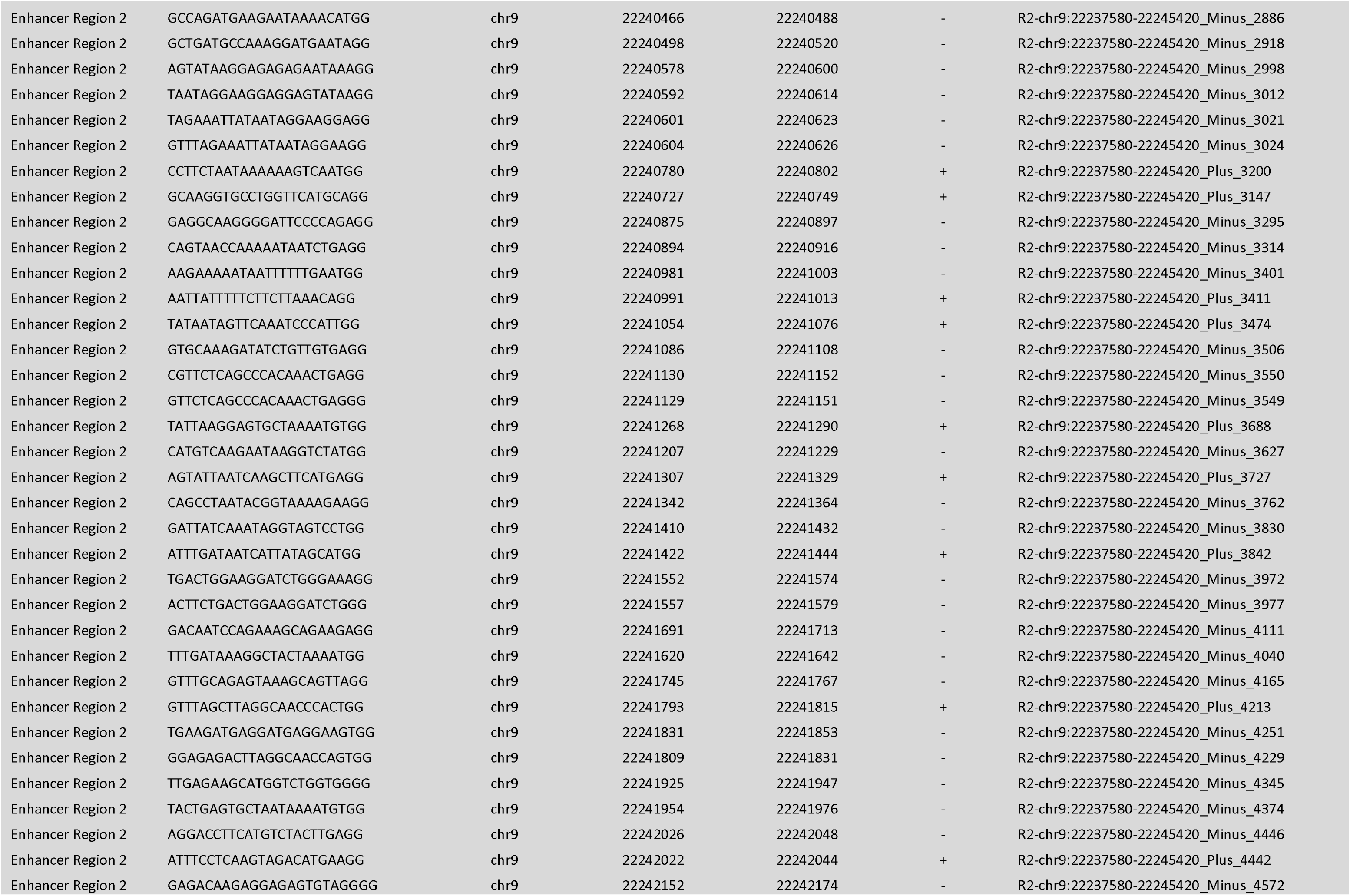
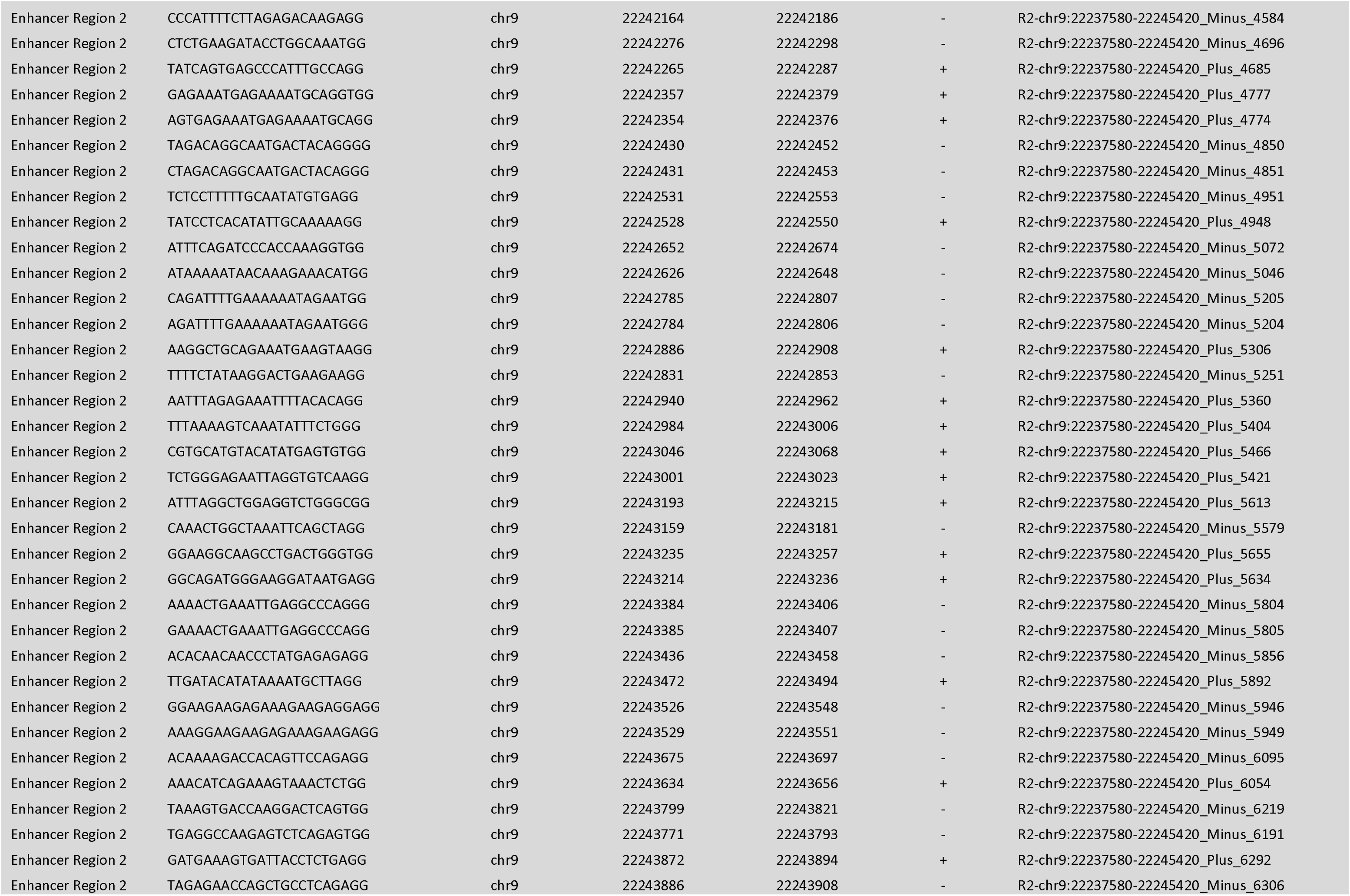
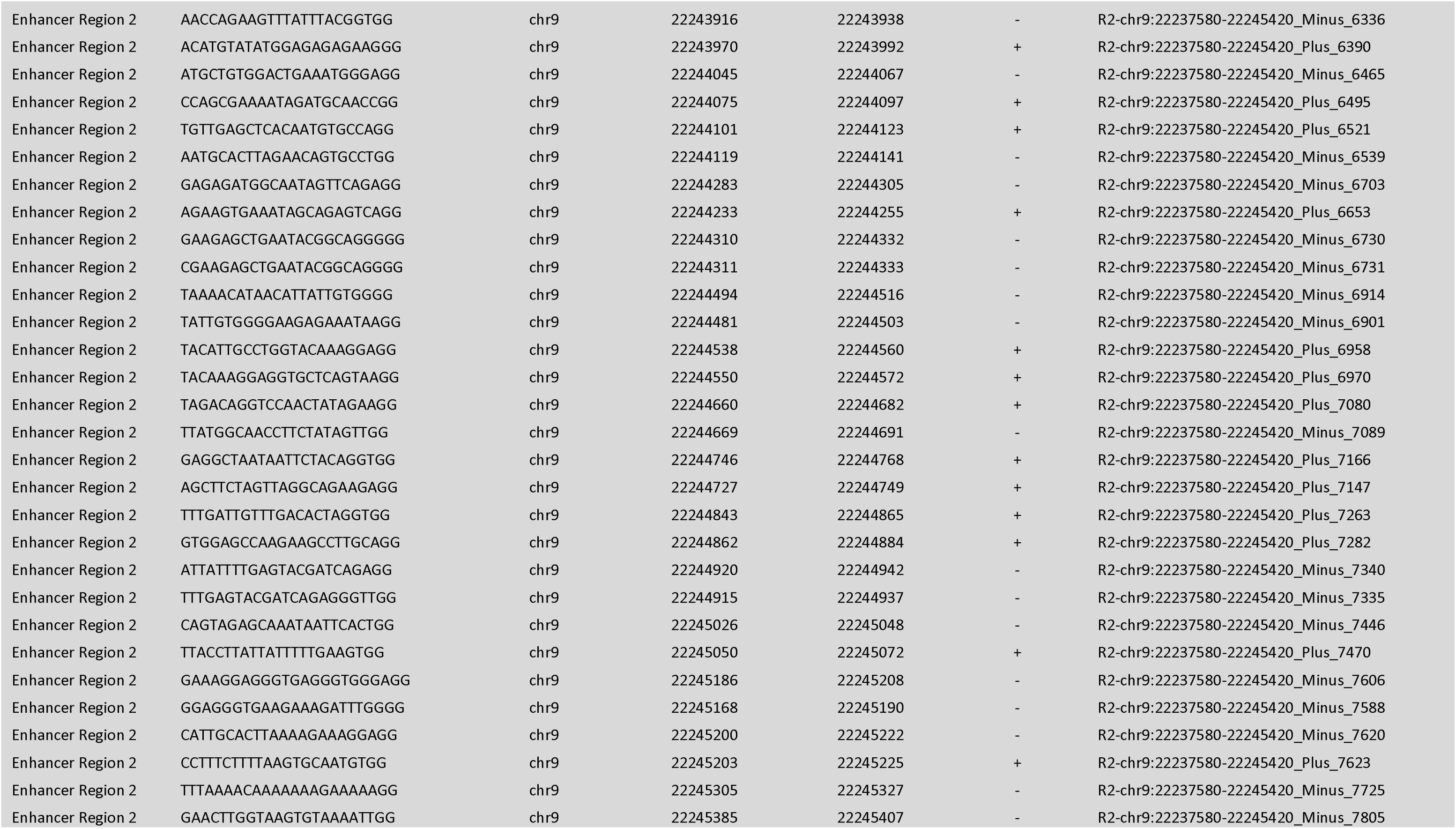
sgRNA sequences and coordinates (hg19)

## Data Availability

The BAM files are available from the European Genome-phenome Archive (https://ega-archive.org/) under accession number EGAS50000001496.

## Acknowledgements

We thank the patients and families who participated in this study. We thank Judith Symmons and Hayley Hamilton for their work on the QFMP, the AMFS Investigators for their work on the AMFS cohort and C. Daniela Robles-Espinoza for her work on the British cohorts. This work was supported by NHMRC (1093017, 1117663), Highlands and Islands Enterprise (HMS 9353763), and the Intramural Research Program of the National Institutes of Health (NIH). The contributions of the NIH authors are considered Work of the United States Government. The findings and conclusions presented in this paper are those of the authors and do not necessarily reflect the views of the NIH or the U.S. Department of Health and Human Services. KB was supported by Cure Cancer Australia; VN by an Australian Government Research Training Programme (RTP) Scholarship; CBR by a European Social Fund PhD studentship; and AC by NHMRC Investigator Grant (2008454). We also thank Buck Off Melanoma for their support.

## Author contributions

Conceptualisation: P.A.J., D.J.A., K.M.B., N.K.H. and A.L.P.; Data curation: P.A.J., J.M.P., E.A.H., H.S., L.C., M.Ho., T.D.B., J.N.-B., G.J.M., A.E.C., D.J.A., N.K.H. and A.L.P.; Formal analysis: P.A.J., V.N., M.X., J.L.S., R.H., M.Ha., S.H., P.Y.C., A.S., S.P., A.D., R.T., R.C., M.H.L., C.B.R., C.M. and A.L.P.; Funding acquisition: D.J.A., K.M.B., N.K.H. and A.L.P.; Investigation: P.A.J., K.B., J.M.P., V.N., M.X., J.L.S., R.H., E.A.H., M.Ha., S.H., P.Y.C., A.S., S.P., A.D., H.S., M.H.L., M.Ho., C.B.R., C.M., T.D.B., J.N.-B., G.J.M., A.E.C. and A.L.P.; Methodology: P.A.J., K.B., M.X., S.H., R.C., C.B.R., D.J.A., K.M.B. and A.L.P.; Project administration: P.A.J., N.K.H. and A.L.P.; Resources: P.A.J., J.M.P., E.A.H., R.T., R.C., H.S., M.H.L., L.C., M.Ho., T.D.B., J.N.-B., G.J.M., A.E.C., D.J.A., N.K.H. and A.L.P.; Software: P.A.J., M.H.L. and D.J.A.; Supervision: P.A.J., M.X., J.L.S., R.H., D.J.A., K.M.B., N.K.H. and A.L.P.; Validation: P.A.J., V.N., M.X., M.Ha., S.H., P.Y.C., A.S., M.H.L., C.B.R., C.M. and A.L.P.; Visualisation: P.A.J., J.M.P., M.X., M.H.L., K.M.B. and A.L.P.; Writing - original draft: P.A.J., M.X., K.M.B., N.K.H. and A.L.P.; Writing - review and editing: P.A.J., K.B., J.M.P., V.N., J.L.S., E.A.H., M.Ha., P.Y.C., A.S., S.P., A.D., H.S., M.H.L., L.C., M.Ho., C.B.R., C.M., T.D.B., J.N.-B., G.J.M., A.E.C., D.J.A., N.K.H. and A.L.P.

